# Improving Hospital Readmission Prediction using Individualized Utility Analysis

**DOI:** 10.1101/2020.07.26.20156943

**Authors:** Michael Ko, Emma Chen, Pranav Rajpurkar, Ashwin Agrawal, Anand Avati, Andrew Ng, Sanjay Basu, Nigam H. Shah

**Affiliations:** Department of Computer Science, Stanford University, CA, USA; Center for Primary Care, Harvard Medical School, MA, USA; Center for Biomedical Informatics Research, Stanford University, Stanford, CA, USA

## Abstract

**Importance:** Machine learning (ML) models for allocating readmission-mitigating interventions are typically selected according to their discriminative ability, which may not necessarily translate into utility in allocation of resources.

**Objective:** To determine whether ML models for allocating readmission-mitigating interventions are ranked differently based on their overall utility and their discriminative ability.

**Design:** A retrospective analysis of ML models using claims data acquired from the Optum Clinformatics Data Mart.

**Setting:** Health plan claims from all 50 states for commercially-insured individuals.

**Participants:** 513,495 patients who were admitted as inpatients over the period January 2016 through January 2017.

**Main Outcomes and Measures:** Maximum utility achieved by three machine learning models for allocating readmission-mitigating interventions, determined using cost accrued in the 90 days post-discharge of an index admission and estimated counterfactual cost. Data were analyzed between April 2019 and March 2020.

**Results:** The study sample consisted of 513,495 patients (mean [SD] age 69 [19] years; 294,895 [57%] Female) mean 90 day cost of $11,552 for the study period. Allocating readmission-mitigating interventions based on a LightGBM model trained to predict readmissions achieved a maximum utility of $-12,645 per patient, and an AUC of 0.74 (95% CI 0.74, 0.75); allocating interventions based on a model trained to predict cost as a proxy achieved a higher maximum utility of $-12,472 per patient, and an AUC of 0.63 (95% CI 0.62, 0.63). A hybrid model combining both intervention strategies achieved a maximum utility of $-12,472, and an AUC of 0.71 (95% CI 0.71, 0.71), comparable with the best models on either metric.

**Conclusion and Relevance:** We demonstrate that machine learning models may be ranked differently based on overall utility and discriminative ability. Machine learning models for allocation of limited health resources should consider directly optimizing for utility.

**Key points:** *Question:* Do machine learning models for allocating readmission-mitigating interventions rank differently based on overall utility and discriminative performance?

*Finding:* A machine learning model predicting a patient’s future cost of care was able to achieve higher utility than a readmission risk prediction model, even though it had a lower discriminative performance in predicting readmissions.

*Meaning:* Our study suggests that machine learning models guiding allocation of limited health resources may consider evaluating and optimizing on utility.

## Introduction

Machine learning–based models for predicting a future health state are now common, however, evidence that the use of these models to guide interventions has improved patient outcomes is lacking^1^. For example, despite a number of machine learning models for allocating readmission mitigating interventions demonstrating high accuracy, there is scant evidence that the guidance by these models improves care and patient outcomes^2^. A key reason for this gap is that typical measures of predictive performance, such as discrimination (the ability to differentiate people at higher risk of having an event from those at lower risk) and calibration (the extend to which the values predicted agree with the observed absolute risk among a group of population)^3^, do not necessarily reflect clinical usefulness^1^. Hospital readmissions are a well known example of this disconnect. Unplanned readmissions have a high financial burden for hospitals^4^, are associated with adverse patient outcomes^5^, and are reflections of low quality of care^6^. The Medicare Payment Advisory Commission (MedPAC) has estimated that 12% of readmissions are potentially avoidable, and has estimated the potential cost-savings at $1 billion^7^.

In this study, we investigated how ML strategies for allocating readmission-mitigating interventions are ranked based on utility. Utility is measured by taking into account the dollar value costs of an intervention allocation and treatments, along with estimates of future individual patient expenses. We examined whether higher discriminative ability necessarily translated into improved utility with three models that we developed: one to directly predict 30-day readmission using claims data, another to predict 90-day cost as a proxy for the task of readmission prediction, and a hybrid model that incorporated the predictions of the 2 models. We compared the performance of all three models on discriminative ability measured by AUC and utility measured by individualized cost saving estimates.

## Methods

### Ethics

The Stanford University administrative panel for the Protection of Human Subjects approved this study.

### Data

Claims data were acquired from the Optum Clinformatics Data Mart, which collects administrative health claims for commercially insured members nationally. Claims data were verified, adjudicated, adjusted with a standard pricing methodology to account for differences in pricing across health plans and provider contracts, and de-identified prior to inclusion in the Data Mart dataset. For this analysis, data are health plan claims from all 50 states for commercially-insured individuals who were admitted as inpatients over the period January 2016 through January 2017. The data include demographics (age, sex from enrollment applications) and all medical claims data, including inpatient visits, International Classification of Disease series 10 diagnostic codes, and payments. The data were split into a training set (80%) used for model development and selection, and a test set (20%) used for model testing.

### Outcome

The primary outcome in this study was all-cause unplanned readmission within 30 days of discharge. A readmission was defined as a subsequent hospitalization following an eligible index admission. An index admission was eligible if the following criteria were met: (1) patient was enrolled for 12 consecutive months prior to discharge and 90 continuous days post-discharge, (2) patient was discharged alive, (3) patient did not leave against medical advice, (4) patient was not transferred to another acute care hospital setting, (5) patient’s primary diagnosis was not one among psychiatric disorders, cancer treatments or rehabilitation care, (6) patient’s previous index admission was not within 30 days prior to discharge. Eligible index admissions were considered to have a subsequent readmission if the patient had an unplanned admission within 30 days of discharge. An admission was defined as “planned” and not part of the readmission measure if it contained procedures from a set of pre-specified planned procedure codes, i.e. organ transplants or chemotherapy, and did not include acute diagnosis codes for potentially planned procedures as described by Planned Readmission Algorithm (Version 4.0, 2019)^8,9^. All other readmissions were considered as unplanned, regardless of cause. The outcome used in this study was consistent with the methodology used in the U.S. Centers for Medicare & Medicaid Services (CMS) definitions, with the exception that we required 90 continuous days of enrollment post-discharge, rather than 30 days, for the purposes of our cost analysis. The details of the labelling and cohort exclusions procedure adapted can be viewed in Appendix D & E of CMS 2019 Measures Update: HWR^9^.

The secondary outcome in this study was the cost accrued in the 90 days post-discharge of an index admission, herein referred to as post-discharge cost. The accrued cost was defined as total standardized gross payments (not charges) to all providers and facilities, computed for each patient index admission by summing standardized costs (in US Dollars) over 90 days, including post year claims corrections, and including zero spending among enrolled individuals without medical claims.

### Model Development and Validation

We developed machine learning models using gradient boosted decision trees (GBDTs) to predict readmission risk (readmission model), and post-discharge cost (cost model). GBDTs employ decision trees to capture nonlinear relationships in data that traditional linear models are unable to capture, and can handle mixes of categorical and continuous covariates^10^. The training procedure for GBDTs involves the construction of a sequence of decision trees such that each tree learns from the errors of the prior tree to iteratively improve predictions^11^.

We trained the readmission and cost models on an identical set of features extracted for every index admission. We included demographics (age, sex) associated with the patient, and diagnostic and procedure codes encoded as aggregated counts of Clinical Classification Software categories^12^ from 12 months prior to discharge. In addition, the following features included in the HOSPITAL Score^13^ were computed: (1) number of procedures during hospital stay, (2) number of admissions in the previous year, (3) number of hospital stays for 5 or more days, and (4) type of index admission (urgent or elective). All features included in the LACE Index^14^ were also computed: (1) length of stay, (2) whether the admission was acute, (3) Charlson Comorbidity Index^15^, and (4) number of ED visits prior to the index admission. A total of 539 features were used.

We used 3-fold cross-validation on the training data subset to select the hyper-parameters for the models, including the number of trees, the maximum depth of each tree, and the required minimum loss reduction to partition leaf nodes based on the cross-validation area under the receiver operating characteristic curve (AUC) and R2 for the readmission model and cost model respectively. Each model was refitted to the full training set using the best parameters determined from 3-fold cross validation.

The readmission model, cost models and an additional hybrid model, were all built for the task of unplanned readmission prediction. The hybrid model multiplied the output of the readmission model with the output of the cost model. We evaluated the discriminative performance of these models. We reported the AUC of all these models on the test set along with 95% confidence intervals computed using the nonparametric bootstrap method with 1,000 bootstrap replicates.

### Individualized Utility Analysis

We used an approach, which we refer to as Individualized Utility Analysis (IUA), for comparing predictive models. IUA estimates the cost, in dollars, of allocating interventions for reducing readmission risk based on the reduction in the 90-day cost. To avoid the problem of heterogeneity in outcomes and costs across patients^16^, IUA estimates the cost on an individual level. Although we present the example using dollar cost, other units such as estimates of disability adjusted life years can also be used for such utility calculations.

IUA constructs a patient-outcome cost matrix using observed patient cost, estimated counterfactual patient cost, treatment cost and treatment efficacy. IUA makes the following assumptions: (1) treatment cost and efficacy is fixed across patients (2) the estimated counterfactual cost is a good estimate of the cost incurred for a patient, had they received an effective intervention.

The calculations for obtaining the expected future cost for each scenario are summarized in Figure 1. The expected 90-day cost outcome of different decisions are estimated for each patient based on treatments assigned by a given predictive model. A predictive model can assign ‘treatment’ or ‘no treatment’ for every patient. If treatment is not assigned, the future cost will be the actual cost in the dataset for both readmitted and not-readmitted patients. If treatment is assigned, there are two scenarios. There is one subgroup who will get readmitted, and for them, giving a treatment is effective with probability p. For the other subgroup who will not get readmitted, assigning a treatment to them is assumed to be harmless (i.e. a treatment will not lead to readmission). If treatment is assigned to the not-readmitted subgroup, their future cost is the actual cost plus the treatment cost. If treatment is assigned to the readmitted subgroup, the cost is defined to be the weighted average of a counterfactual cost (cost if a readmitted patient did not have that readmission) and their observed cost with the efficacy of the treatment serving as the weight. We assume a linear u-curve (risk neutral assumption) for the utility.

**Figure 1:**
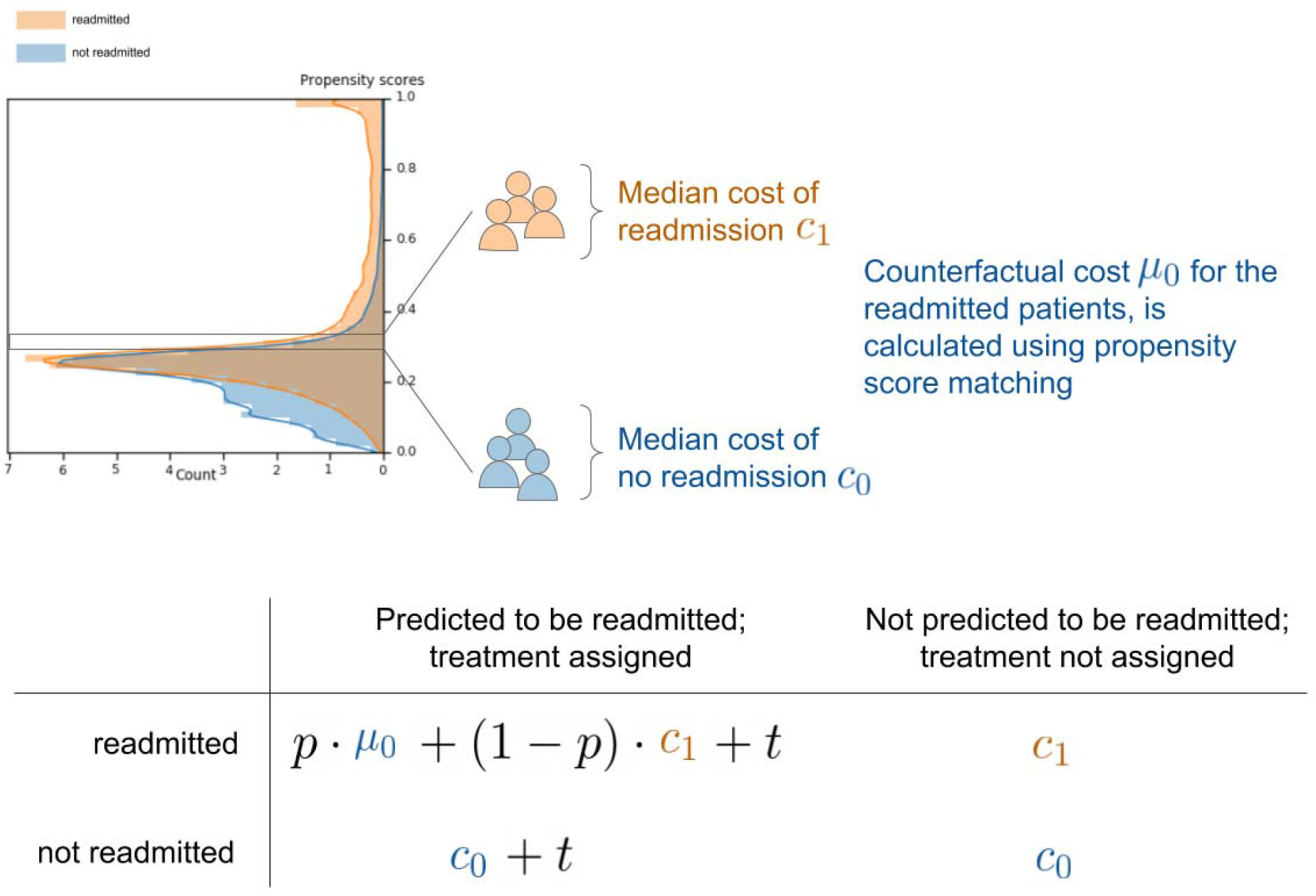
Computation of an individual’s cost estimate. Propensity scores were calculated using an L1 regularized logistic regression model for 90-day cost. The distribution of propensity scores showed sufficient overlap between readmitted and not readmitted population in the dataset (top left). We used propensity score matching procedure to assign patients to one of 20 equal width propensity score bins from 0.0 to 0.6. Within each bin, the median of the non-admitted patient group was used to impute the counterfactual costs for the readmitted patients, had they not had the readmission (μ0). Upon assigning a treatment based on one of the machine learning derived models, the 90-day cost of the patient’s care is calculated as follows: *If treatment is not assigned*, the future cost will be the observed cost in the dataset for both readmitted (C1) and not readmitted patients (C0). *If treatment is assigned*, there are two scenarios. If treatment was assigned to the not readmitted subgroup, the future cost is the actual cost (C0) plus the cost of the treatment (t). The effect of the treatment on them is assumed to be harmless, meaning a reamission will not occur due to the treatment. If treatment was assigned to the readmitted subgroup, the future cost is defined to be the weighted average of their counterfactual cost (μ0) and their observed cost (C1) with the efficacy of the treatment (p) serving as the weight.

We used propensity score matching (PSM)^17^ to estimate counterfactual cost using similarity between sets of treatment and control groups in probability of readmission. Propensity scores were obtained from an L1 regularized logistic regression model trained to predict 90-day cost using an identical feature set and hyperparameter selection procedure as the readmission and cost models. The matching procedure assigned patients to one of 20 equal width propensity score bins from 0.0 to 0.6. Within each bin, the median of the non-admitted patient group was used to impute the counterfactual costs for the readmitted patients (i.e. their cost if they would not have a readmission). We evaluated the goodness of fit of the logistic regression model on the test set using R2 and mean absolute error, with its 95% confidence intervals computed using the non-parametric bootstrap with 1000 bootstrap replicates.

The training dataset was split into non-overlapping subsets, with 12.5% used to train the propensity score model, and 87.5% used to train the machine learning models.

### Feature Importances

We quantified the impact of each input feature on the readmission and cost models. The SHAP (SHapley Additive exPlanations) method was used. The method explains prediction by allocating credit among the input features; feature credit is calculated using Shapley Values^18^, as the change in the expected value of the model’s prediction of improvement for a symptom when a feature is observed versus unknown. To uncover clinically important features that were globally predictive of the readmissions outcome, the Shapley values for features on individual predictions were aggregated and reported along with their averaged absolute Shapley contributions as a percent of the contributions of all the features.

## Results

The final dataset consisted of 513,495 patients, divided into a training set of 410,796 patients, a test set of 102,699 patients. There was no patient overlap between the two sets.

Overall, the average number of index admissions per patient was 1.18. The prevalence of readmission was 25.41%, and the mean 90-day cost was $11,552. Table 1 details the patient, admission, and outcome characteristics in the dataset.

**Table 1:** Summary statistics of the training set, test set, and overall dataset.

| Characteristics | Training Set | Test Set | Overall |
| --- | --- | --- | --- |
| Total Members | 410,796 | 102,699 | 513,495 |
| Total Female | 235,836 (57.41%) | 59,059 (57.51%) | 294,895 (57.43%) |
| Age, mean (std) | 69.11 (18.79) | 69.15 (18.76) | 69.12 (18.79) |
| Total Index Admission | 484,609 | 121,265 | 605,874 |
| Readmission Rate | 123,405 (25.46%) | 30,574 (25.21%) | 153,979 (25.41%) |
| Mean 90-day cost | \$ 11,556 | \$ 11,538 | \$ 11,552 |
| 25<sup>th</sup> Quantile 90-day cost | \$ 415 | \$ 418 | \$ 415 |
| 50<sup>th</sup> Quantile 90-day cost | \$ 2,521 | \$ 2,521 | \$ 2,521 |
| 75<sup>th</sup> Quantile 90-day cost | \$ 11,556 | \$ 11,538 | \$ 11,552 |

### Model validation

On the task of predicting unplanned readmissions, the readmission model achieved an AUC of 0.74 (95% CI 0.74, 0.75) on the test set; the cost model achieved an AUC of 0.63 (95% CI 0.62, 0.63); the hybrid model achieved an AUC of 0.71 (95% CI 0.71, 0.71). On the task of predicting cost, the cost model achieved an R2 of 0.21 (95% CI 0.19, 0.22) and a mean absolute error of $13,955 (95% CI $13,815, $14,095). The receiver operating characteristic (ROC) curves for each of the models are shown in Figure 2.

**Figure 2:**
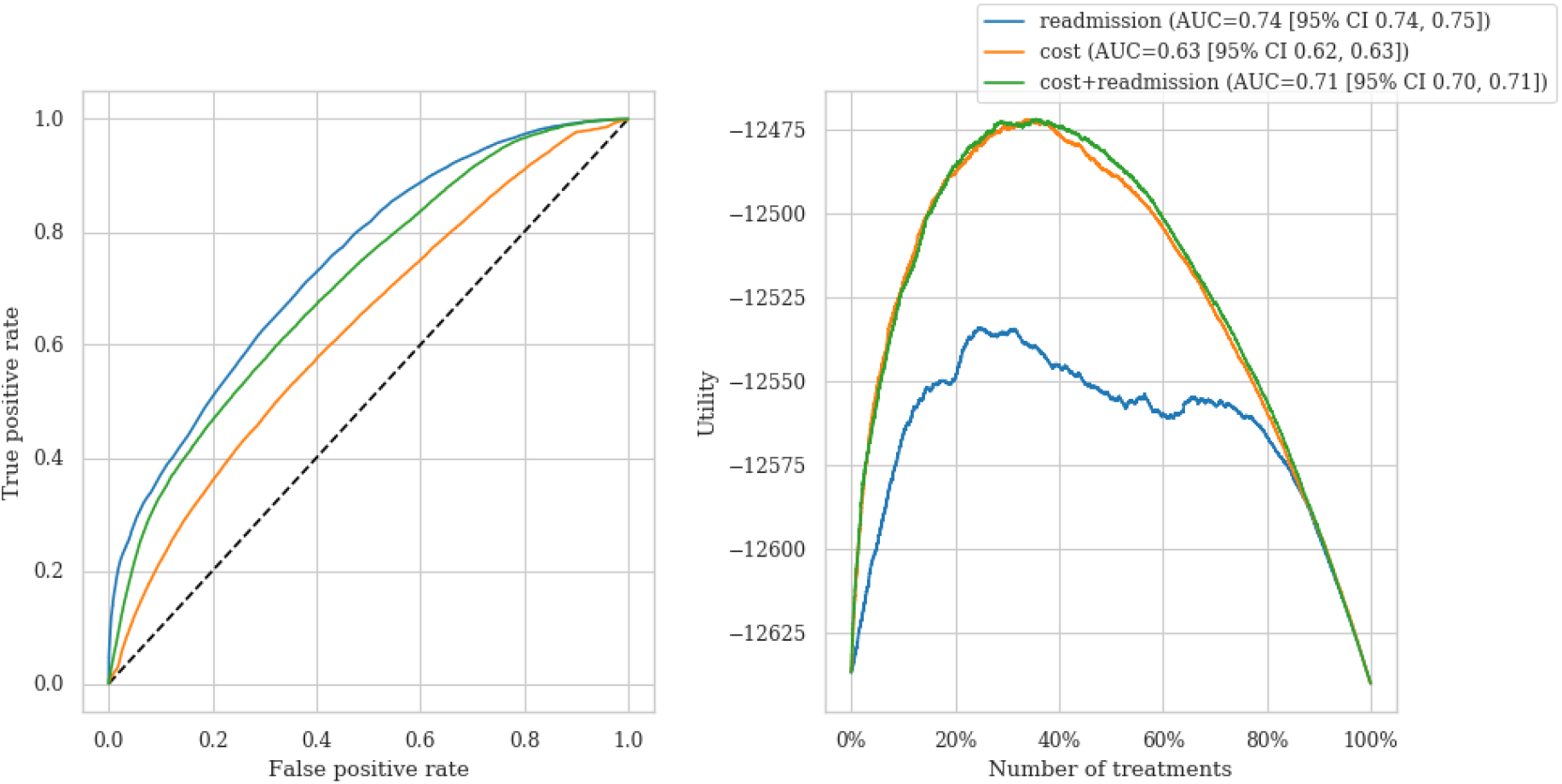
The left panel shows the ROC curve for three models for assigning treatments to prevent a readmission: a readmission model (AUC = 0.74), which assigns treatment according to readmission probability, a cost model (AUC = 0.63), which assigns treatment according to the cost of patient, and a hybrid cost + readmission (AUC = 0.71), which assigns treatment considering the dot product of cost and readmission probability. The right panel shows the utility realized in terms of the average savings per patient given a certain number of treatments provided. With a fixed treatment cost of $500 and efficacy of 10%, the readmission model had a maximum utility of $-12,645 per patient when 24.5% of the patient population was treated, the cost model had a maximum utility of $-12,472 per patient when 33.6% of the patient population was treated, and the hybrid model had a maximum utility of $-12,472 when 35.5% of the patient population was treated.

### Individualized Utility Analysis

As computed by the IUA approach using a fixed treatment cost of $500 and efficacy of 10%, the readmission model had a maximum utility of $-12,534 per patient when 24.5% of the patient population was treated, the cost model had a maximum utility of $-12,472 per patient when 33.6% of the patient population was treated, and the hybrid model had a maximum utility of $-12,472 when 35.5% of the patient population was treated. Both the cost and hybrid models’ maximum utilities per patient were significantly greater than that of the readmission model using bootstrapping and bonferroni correction (p<.001). The propensity score model used in IUA had an AUC of 0.691 (95% CI 0.688, 0.695), a calibration slope of 0.945 (95% 0.931, 0.957) and a calibration intercept of 0.0124 (95% 0.0085, 0.016).

### Feature Importances

The most important features for the readmission prediction model were the number of hospital admissions (higher value increased risk), Osteoarthritis (higher value decreased risk) and Admission type (higher value decreased risk) with mean contributions of 18.5%, 10.5% and 11.0%.

For the cost model, the most important features for were Hemodialysis (higher value decreased cost), length of stays in the hospitals (higher value increased cost) and the Comorbidity index of the LACE model (higher value decreased cost) with mean contributions of 17.0%, 10.5% and 8.0%. A summary of the top 10 most important features for both the readmission and cost models is detailed in the Figure 3.

**Figure 3:**
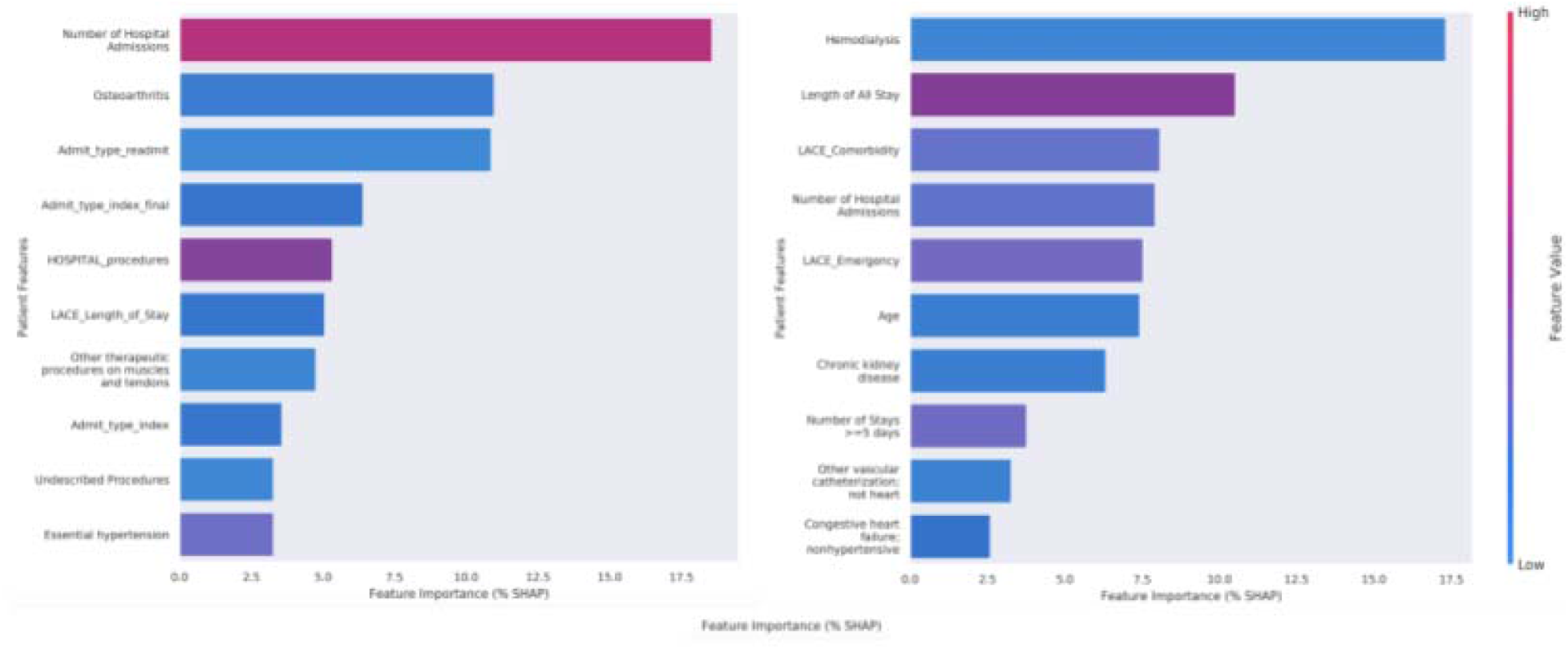
Important features for the readmission model (left) and cost model (right). The impact of each feature on the discrimination ability of the readmission and cost models was quantified using the SHAP (SHapley Additive exPlanations) method, which explains prediction by allocating credit among the input features. Feature importances on individual predictions were aggregated and reported along with their averaged absolute Shapley contributions as a percent of the contributions of all the features.

## Discussion

In this study, we assessed the utility of machine learning models on the task of predicting unplanned readmissions. In this approach, we took into account the dollar value costs of an intervention allocation and treatments, along with estimates of future individual patient expenses. Our study led to three main findings. First, although a model trained to predict readmissions achieved a higher AUC than a model trained to predict cost (as a proxy for readmission), the model trained to predict cost achieved a higher utility if interventions to prevent readmissions were allocated based on the ranking of patients produced by the cost model. Second, a hybrid model that combined the predictions of the readmission model with the cost model achieved comparable utility and higher AUC to the cost model. Third, we found--as numerous prior studies have--that the number of prior hospital admissions was the most important feature for the readmission model^19^, and the fourth most important feature for the cost model.

Our results suggest that classification or prediction models with higher discriminative performance may not necessarily give higher utility. In healthcare, there is always a capacity constraint (either monetary or staffing) which induces a hard limit on the number of actions that can be taken in response to a model’s ranking of patients at risk of an outcome such as readmission^20,21^. The intent underlying most risk-scoring^22^ (from readmissions^23^, to cost^24^, to mortality^25^) is to allocate interventions to those in most need. The factors constituting utility can be different in different healthcare settings and problem formulations. As a result, utility per unit cost (or the budget) becomes one objective measure for evaluating clinical risk scoring.

Therefore we use cost as the measure of utility for the readmission prediction task. We acknowledge that “cost” is an imperfect proxy for need across patient groups as outlined by Obermeyer et al^24^ and that caution needs to be exercised in defining the outcome for which a prediction model is built^24^. However, within a homogenous patient group where label-bias can be avoided, incorporating cost information directly into model building provides a much tighter alignment with the end goal--which is allocation of limited resources to derive the highest value from interventions.

Our proposed approach for analyzing individualized utility poses several advantages over prior literature studying 30-day readmission^14^. Our approach uses a model evaluation that focuses on utility obtained through taking actions based on predictions whilst most 30-day readmission work has focused on classification metrics such as AUC that do not account for the cost implications of alternative actions^23^. Although a case study of reducing readmissions for heart failure done by Bayati et al., has demonstrated that improvements in prediction quality can improve utility^2^, a key difference is that the prior work used the average cost of readmission in calculating utility, which may not translate into an individual-level analysis^16^. We found that 90 day costs have a high variability by patient and our individualized utility analysis approach appropriately captures this complexity. Prior work that uses global estimates to measure net benefit are also limited in assuming that averages are generalizable to everyone^2,26^.

### Limitations

This study has several important limitations. First, utility was defined as the dollar value with a linear U-Curve, and may be redefined to incorporate measures such as quality of life, number of actions a care team takes, essential resources consumed with a nonlinear U-curve. Second, our analysis is based on retrospective data. Rigorous validation of our findings using individualized utility analysis for estimating the utility of 30-day readmission models would require evaluation in a prospective setting. Third, we do not investigate the performance of models when explicitly performing model selection based on our IUA method; we expect that training procedures that directly optimize and select based on the utility may be a fruitful avenue for further exploration.

## Conclusion

Machine learning models often focus on predicting the probability of occurrence of an outcome, in an effort to allocate limited health resources to get higher value for the care delivered. We show that models for intervention allocation can be made more useful by jointly ranking on individual utility and discriminative ability. Machine learning models for allocation of limited health resources should consider directly optimizing for utility.

## Data Availability

NA

